# Tumor–blood TCRβ repertoire architecture in endometrial cancer: abundance-dependent peripheral detection and intratumoral enrichment of shared clonotypes

**DOI:** 10.64898/2026.06.08.26355191

**Authors:** Ilenia Aversa, Antonio Abatino, Aida Isabello, Raffaella Gallo, Lisa Isdraele Romano, Terry Straface, Carmela De Marco, Maria Teresa De Angelis, Francesco Maria Zullo, Maurizio Guida, Gabriele Saccone, Claudia Casella, Giuseppe Fiume, Roberta Venturella, Francesco Costanzo, Giovanni Cuda, Giuseppe Viglietto, Fulvio Zullo, Camillo Palmieri

**Affiliations:** Department of Experimental and Clinical Medicine, University Magna Grecia of Catanzaro; 88100 Catanzaro, Italy; UOC Biochimica Clinica, AOU Renato Dulbecco, Catanzaro, Italy; UOC Ginecologia e Ostetricia, AOU Renato Dulbecco, Catanzaro, Italy; Interdepartmental Centre of Services, Magna Graecia University of Catanzaro, Catanzaro, Italy; Department of Neuroscience, Reproductive Sciences and Dentistry, University of Naples Federico II, Naples, Italy

## Abstract

Endometrial cancer exhibits substantial molecular and immune heterogeneity that is incompletely captured by established genomic and histopathological descriptors. We investigated whether paired tumor–blood T-cell receptor β (TCRβ) profiling could provide a complementary view of adaptive immune organization by jointly resolving intratumoral repertoire structure, abundance-dependent peripheral detectability, and genomic correlates.

TCRβ repertoires were profiled in paired tumor biopsies and peripheral-blood mononuclear cells from 13 patients with molecularly characterized endometrial cancer. Clonotypes were defined by exact CDR3 amino-acid identity, and repertoire differences were evaluated at observed sequencing depth and under multiple depth- and coverage-standardization procedures. Tumor–blood sharing was assessed after 500 pair-specific matched-depth resamplings.

As expected, tumor biopsies contained fewer clonotypes than matched blood (13/13 patients); this difference was preserved across depth- and coverage-standardization procedures, indicating that this compartmental difference was not explained by sequencing depth alone. Peripheral detectability increased with intratumoral abundance, with the odds of blood detection rising 2.44-fold per tenfold increase in tumor frequency. Among shared clonotypes, 66.7% were at least twofold more frequent in tumor versus 8.7% in blood, consistent with preferential intratumoral enrichment. In a prespecified 16-test immune–genomic analysis, higher tumor mutational burden was strongly associated with lower intratumoral Hill D1 (Spearman ρ=−0.833; BH q=0.0115; max-|ρ| FWER P=0.0088), with consistent leave-one-subject-out direction and persistence after excluding hypermutated tumors. No histopathological endpoint showed a multiplicity-corrected association.

Together, these findings define a quantitative tumor–blood TCRβ architecture in endometrial cancer and support repertoire profiling as a complementary functional layer for future immunogenomic stratification and validation.

**Significance:** Paired tumor and blood TCRβ profiling revealed that clonotypes abundant in endometrial tumors were more often detectable in blood and that shared clonotypes were preferentially enriched in tumor tissue. Intratumoral effective diversity was lower with increasing tumor mutational burden, supporting TCR repertoire profiling as a complementary layer for immunogenomic characterization.

## Introduction

Endometrial cancer is one of the most common gynaecologic malignancies worldwide, with more than 69,000 new cases annually in the United States alone^1^, and is characterized by substantial molecular and immunological heterogeneity. The Cancer Genome Atlas (TCGA) classification identified four biologically distinct groups, POLymerase Epsilon (POLE) ultramutated, microsatellite instability (MSI)/hypermutated, copy-number low, and copy-number high, which differ in genomic architecture, prognosis, and immune contexture^2^. POLE-mutated and mismatch-repair-deficient (MMRd) tumors typically show high tumor mutational burden (TMB)^3^, increased predicted neoantigen load, and prominent lymphocytic infiltration^4,5^, linking tumor genomics to adaptive immunity. Genomic features alone, however, do not fully account for the organization of the antitumor immune response: immune phenotype and response to immune-checkpoint blockade vary substantially among tumors of the same molecular class^5,6^, T-cell recognition also depends on neoantigen clonality, intratumoral heterogeneity, and antigen presentation^7,8^, and tumors within one molecular subgroup can differ markedly in the extent and spatial pattern of immune infiltration^9–11^. T-cell receptor (TCR) repertoire profiling offers a complementary view. The composition and abundance distribution of TCR clonotypes reflect the population structure of the T-cell response, and comparison of tumor and peripheral blood identifies clonotypes represented in both compartments^12^. Such measurements are not direct evidence of tumor-antigen specificity, but across cancer types they have been linked to mutational and neoantigen landscapes, tumor immune states, and response to immune-checkpoint blockade^12,13^.

In endometrial cancer, direct evidence remains limited. An intratumoral TCRβ clonality index was associated with immune gene expression and longer progression-free survival in 32 endometrioid tumors, but blood was not examined^14^. In paired tumor and blood repertoires from 39 patients with gynecologic cancer, eight of them endometrial, clonotypes found at several tumor sites were preferentially represented in blood^15^; tumor-matched clonotypes have also been followed in the blood of patients with endometrial cancer during pembrolizumab treatment^16^ and years after curative treatment of POLE-mutant disease^17^, and single-cell studies across cancer types show that clonotypes expanded in tumors can typically be detected in blood^18^. None of these studies quantified how peripheral detectability varies with a clonotype’s abundance within a single tumor or controlled for sequencing depth and sampling coverage. Whether intratumoral abundance itself governs peripheral detectability in endometrial cancer, and in which direction shared clonotypes are enriched, therefore remains open.

Here, using paired tumor and peripheral-blood TCRβ repertoires from an endometrial-only cohort, we verified that compartmental differences in repertoire breadth were robust to sampling standardization, quantified peripheral detectability as a continuous function of intratumoral clonotype abundance, assessed the direction of the tumor-to-blood abundance asymmetry among shared clonotypes, and tested the association of prespecified repertoire features with tumor genomic and histopathological characteristics.

## Results

### Tumor-biopsy repertoires show reduced observed breadth with preserved organization of dominant clonotypes

We analyzed paired tumor-biopsy and peripheral-blood mononuclear cell (PBMC) TCRβ repertoires from 13 patients with endometrial cancer. Median age was 65 years (IQR, 61–70; range, 54–73). Eleven tumors were classified as endometrioid carcinoma (84.6%), including one reported as endometrioid carcinoma not otherwise specified^19^, and two (15.4%) were clear-cell carcinomas. Among the 11 endometrioid carcinomas, seven were histological grade 1 (G1), three grade 2 (G2), and one grade 3 (G3), according to the International Federation of Gynecology and Obstetrics (FIGO) grading system^20^. Myometrial invasion exceeded 50% of the myometrial thickness in eight of 12 evaluable tumors (66.7%), whereas lymphovascular space invasion (LVSI) was identified in two of 11 evaluable cases (18.2%). Molecular classification assigned two tumors to the POLE-mutant class (POLE p.Pro286Arg and p.Ser459Phe), two to the mismatch-repair-deficient class (MSI-High), two to the p53-abnormal class (TP53 p.Ser241Phe and p.Ala159Asp) and seven to the class with no specific molecular profile (NSMP); one POLE-mutant tumor also carried a subclonal TP53 nonsense mutation and was classified as POLE-mutant according to the hierarchy. The clinicopathological and molecular characteristics of the cohort are summarized in Table 1 and Supplementary Data 1.

**Table 1.** Clinicopathological and molecular characteristics of the cohort. Summary characteristics of the 13 patients with endometrial cancer included in the analysis. Histological grade is reported according to the FIGO system and applies to the 11 endometrioid carcinomas. Myometrial invasion is categorized by the proportion of myometrial thickness involved and was not evaluable in one patient for whom no definitive surgical pathology report was available. LVSI was evaluable in 11 of 13 patients. Molecular classes were assigned with the WHO hierarchical algorithm, with mismatch-repair deficiency defined by MSI-High status and p53-abnormal status by a pathogenic TP53 variant. Patient-level detail is provided in Supplementary Data 1. Abbreviations: EC, endometrioid carcinoma; NSMP, no specific molecular profile.

| Characteristic | Value |
|---|---|
| **Patients, n** | 13 |
| **Age (years), median (IQR; range)** | 65 (61–70; 54–73) |
| **Histotype , Endometrioid, n/N (%)** | 11/13 (84.6%) |
| **Histotype , Clear cell, n/N (%)** | 2/13 (15.4%) |
| **EC with evaluable FIGO grade, n/N** | 11/11 |
| G1 | 7/11 (63.6%) |
| G2 | 3/11 (27.3%) |
| G3 | 1/11 (9.1%) |
| **Myometrial invasion >50%, n/N (%)** | 8/12 (66.7%) |
| **LVSI<sup>c</sup> evaluable, n/N** | 11/13 |
| **LVSI positive, n/N (%)** | 2/11 (18.2%) |
| **Molecular class , POLE-mutant, n/N (%)** | 2/13 (15.4%) |
| **Molecular class , MMRd, n/N (%)** | 2/13 (15.4%) |
| **Molecular class , p53-abnormal, n/N (%)** | 2/13 (15.4%) |
| **Molecular class , NSMP, n/N (%)** | 7/13 (53.8%) |

Across the 13 matched tumor–blood pairs, we characterized 26 TCRβ repertoires. Clonotypes were defined by exact amino acid sequence identity in the complementarity-determining region 3 (CDR3), and their abundance was quantified using read counts. Repertoire architecture was summarized by observed clonotype richness, Hill diversity of order 1 (D_1_; exponential of Shannon entropy) and order 2 (D_2_, inverse Simpson index), and the cumulative relative abundance of the ten most abundant clonotypes (Top-10)^12,21^. Sequencing depth and Good’s coverage were reported as read-based sampling descriptors. Paired compartments were compared with the two-sided Wilcoxon signed-rank test across the 13 pairs. Repertoire-level measurements and sampling diagnostics are provided in Supplementary Data 2.

Biopsy repertoires contained fewer observed clonotypes than their matched PBMC repertoires in all 13 patients (P = 0.0002). Median observed richness was 4,282 clonotypes (IQR, 2,803–5,234) in biopsies, compared with 19,424 (IQR, 13,061–55,500) in PBMC (Figure 1A). Sequencing depth varied by more than two orders of magnitude across biopsies (median 494,200 reads; IQR, 235,971–657,278; range, 6,194–722,423) and 6.6-fold across PBMC samples (median 548,518; IQR, 352,123–707,074; range, 194,746–1,283,973) but did not differ systematically between compartments (P = 0.13). Good’s coverage was nominally higher in biopsies (median 0.9998 versus 0.9979 in PBMC), reflecting the smaller number of observed clonotypes and the correspondingly lower singleton fraction rather than more complete recovery of the underlying repertoire. Depth and coverage are shown in Supplementary Figure S1A–B.

**Figure 1.**
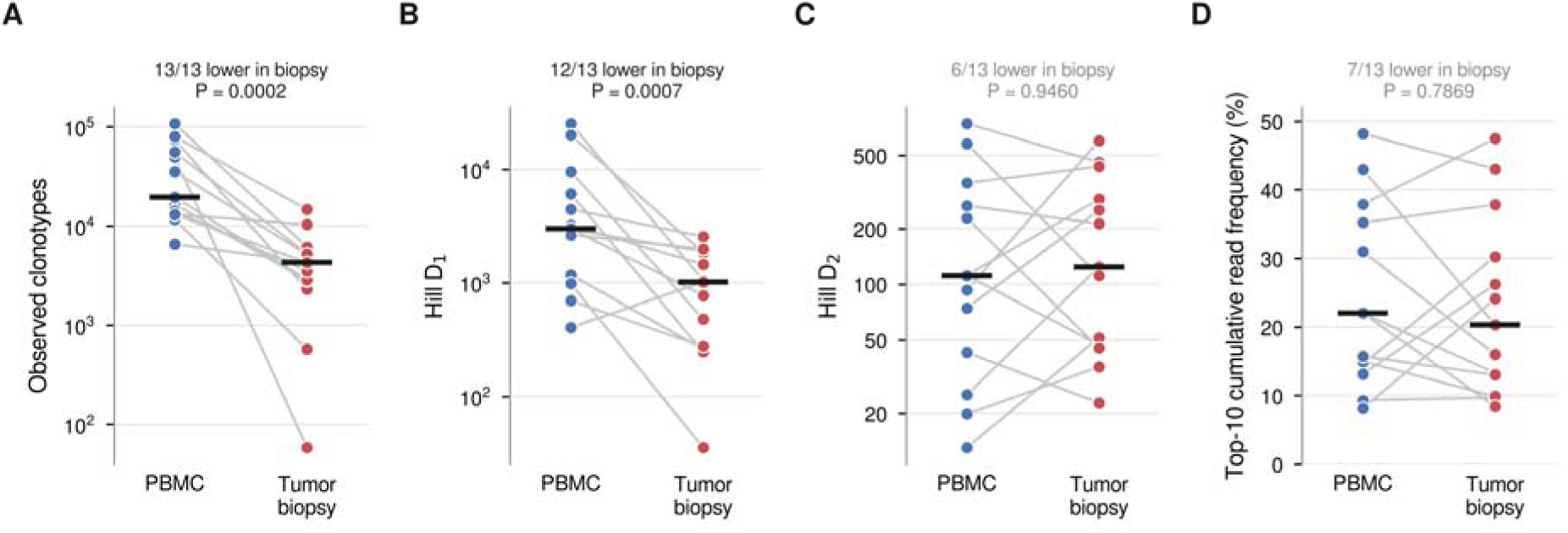
Compartmental architecture of paired tumor-biopsy and peripheral-blood TCRβ repertoires. (A) Observed richness, defined as the number of distinct productive TCRβ clonotypes identified by exact complementarity-determining region 3 (CDR3) amino-acid sequence identity. (B) Hill diversity of order 1 (D_1_), the exponential of Shannon entropy. (C) Hill diversity of order 2 (D_2_), the inverse Simpson concentration, which gives greater weight to abundant clonotypes; both diversity measures are expressed as effective numbers of clonotypes. (D) Cumulative percentage of reads assigned to the ten most abundant clonotypes. Each pair of points joined by a grey line is one patient and horizontal black bars indicate cohort medians; the y-axes in A–C are logarithmic. Directional counts and two-sided Wilcoxon signed-rank P values are given above each panel. All measures were calculated at the observed sequencing depth, without rarefaction; depth- and coverage-standardized versions are shown in Supplementary Figure S1.

The compartmental difference in repertoire breadth persisted after controlling for differences in sequencing depth and sampling coverage. Biopsy richness was lower in all five pairs in which the biopsy yielded more reads than the matched PBMC sample (Supplementary Figure S1C). When the deeper repertoire of each pair was subsampled without replacement to the depth of the shallower member, biopsy richness remained lower in all 13 pairs (P = 0.0002). Rarefying the cohort to a common depth of 63,199 reads (n = 11 pairs, after excluding the two biopsies of lowest depth) and, separately, standardizing all 26 repertoires to a common Good’s coverage of 0.838 gave the same result, with biopsy richness lower in 11 of 11 and 13 of 13 pairs, respectively (P = 0.001 and P = 0.0002). Chao1 richness was likewise lower in biopsies in all 13 pairs (P = 0.0002). Rarefaction curves and depth-standardized paired comparisons are shown in Supplementary Figure S1D–E, and all standardized analyses are tabulated in Supplementary Data 3.

Within biopsies, observed richness was positively associated with sequencing depth (Spearman ρ = 0.75, P = 0.003), whereas no such association was present in PBMC (ρ = −0.08, P = 0.80). This association persisted after rarefaction to a common depth (ρ = 0.73, P = 0.011), indicating that it does not reflect differential sampling effort. It is instead consistent with a shared dependence of both read yield and clonotype number on the amount of T-cell material recovered from the tissue.

The compartmental difference extended to Hill D_1_, the effective number of clonotypes corresponding to Shannon diversity. Median D_1_ was 1,012 (IQR, 275–1,847) in biopsies and 3,002 (IQR, 1,171–6,045) in PBMC, with lower biopsy D_1_ in 12 of 13 patients (P = 0.0007; Figure 1B); the single exception was the pair with the lowest PBMC richness in the cohort (6,566 clonotypes). Lower biopsy D_1_ was recovered in 11 of 13 pairs after per-pair rarefaction (P = 0.002), in 10 of 11 pairs at the common depth of 63,199 reads (P = 0.005), and in 12 of 13 pairs after coverage standardization (P = 0.001). The reduction in observed richness was therefore accompanied by lower Shannon-order effective diversity in most patients, and this held under every standardization applied.

In contrast, measures emphasizing the most abundant clonotypes showed no consistent paired direction and, if anything, pointed towards lower rather than higher dominance in biopsies. Median Hill D_2_ was 124.2 (IQR, 47.1–288.5) in biopsies and 111.3 (IQR, 42.5–265.9) in PBMC, with lower biopsy D_2_ in six pairs and higher D_2_ in seven (P = 0.95; Figure 1C). The ten most abundant clonotypes accounted for a median of 20.3% of biopsy reads (IQR, 13.0–30.2%) and 22.0% of PBMC reads (IQR, 14.5–35.2%), with a higher biopsy Top-10 frequency in six of 13 pairs (P = 0.79; Figure 1D). Neither the Gini coefficient over observed clonotype frequencies (P = 0.69) nor the fraction of clonotypes required to reach 50% of reads (P = 1.00) differed between compartments, and both D_2_ and Top-10 remained without consistent direction under every depth- and coverage-standardized analysis (P ≥ 0.57).

Top-100 cumulative frequency showed a nominal paired difference between compartments, being higher in biopsies in 10 of 13 pairs (median 38.2% versus 30.6%; P = 0.040). Together, these observations locate the compartmental difference in the breadth of the rarer fraction of the repertoire rather than in the organization of its dominant clonotypes.

### Peripheral detection of intratumoral clonotypes scales with their abundance, and shared clonotypes show intratumoral enrichment

Having established differences in repertoire architecture between compartments, we next examined which components of the tumor-biopsy repertoire were also detectable in matched peripheral blood. Sharing was defined by exact TCRβ CDR3 amino-acid identity and evaluated both in the observed repertoires and after 500 pair-specific matched-depth resamplings. Standardized cohort summaries were calculated across patient-specific resampling medians.

After depth matching, a median of 9.2% of biopsy clonotypes were also detected in the corresponding PBMC repertoire (IQR, 6.8–24.5%). These shared clonotypes accounted for a larger fraction of biopsy read abundance, with a median of 20.0% (IQR, 11.3–37.0%; Figure 2A). In the reverse comparison, a median of 1.4% of PBMC clonotypes were detected in the biopsy (IQR, 0.7– 3.1%), representing 24.9% of PBMC read abundance (IQR, 6.0–36.5%). These directional fractions describe sharing relative to the repertoire of each compartment and do not imply a direction of cellular trafficking.

**Figure 2.**
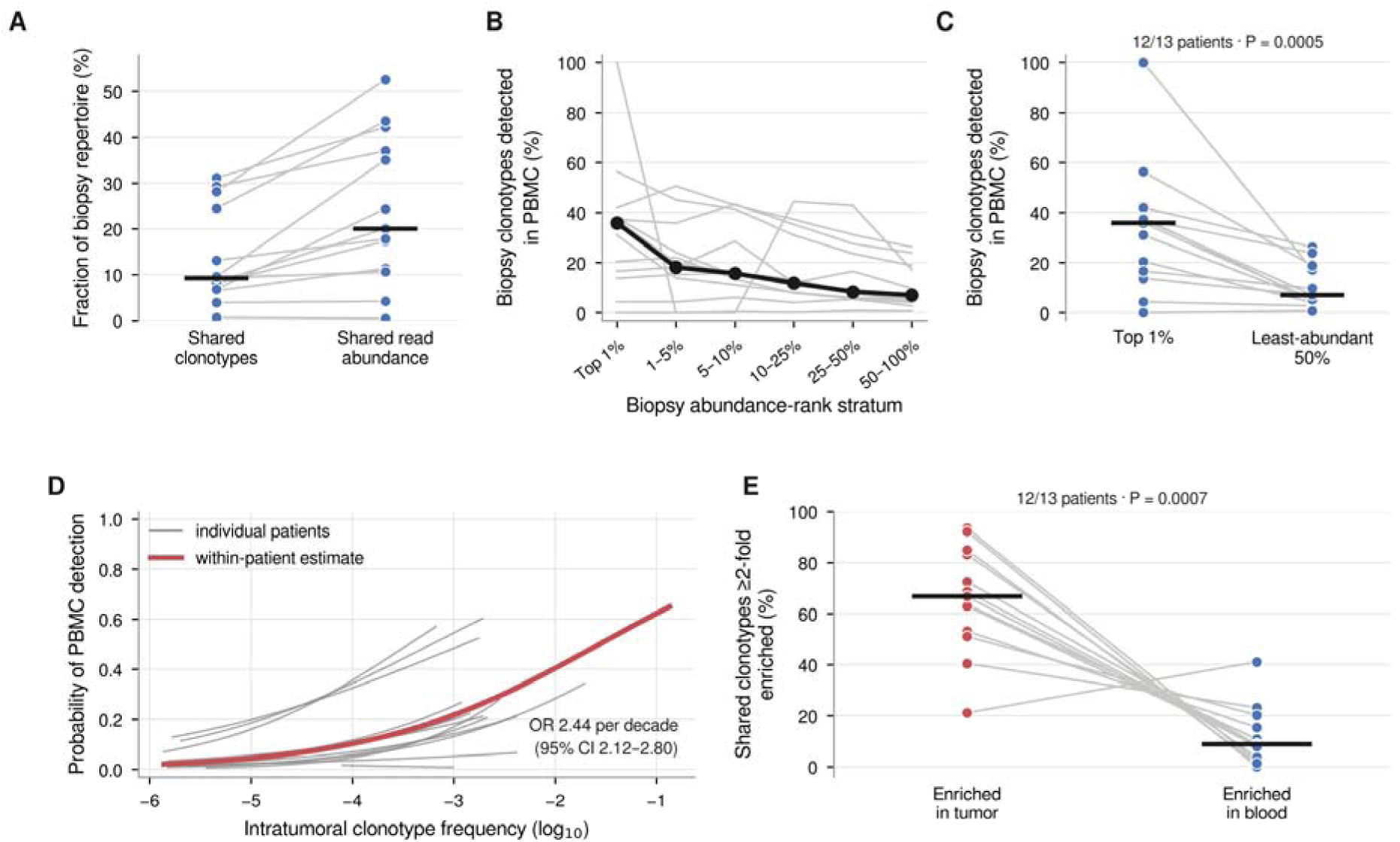
Tumor–blood clonotype sharing and its dependence on intratumoral abundance. (A) Percentage of biopsy clonotypes also detected in matched PBMC, and the percentage of biopsy reads carried by those shared clonotypes. (B) Percentage of biopsy clonotypes detected in PBMC across six non-overlapping abundance-rank strata; thin grey lines are individual patients and the black line with filled circles connects cohort medians. (C) Within-patient comparison between the most abundant 1% and the least-abundant 50% of biopsy clonotypes. (D) Probability that an intratumoral clonotype is also detected in matched PBMC as a function of its intratumoral frequency; thin grey lines are logistic fits within individual patients, each drawn over that patient’s observed frequency range, and the thick line is the within-patient estimate from a single model with patient fixed effects across 62,170 biopsy clonotypes. (E) Among clonotypes detected in both compartments, the percentage at least twofold more frequent in the biopsy and the percentage at least twofold more frequent in blood. Values in A–C are patient-specific medians across 500 pair-specific matched-depth resamplings; grey lines connect measurements from the same patient and horizontal black bars indicate cohort medians.

Despite the contribution of shared clonotypes to read abundance, overall repertoire similarity remained limited. The median matched-depth Jaccard index was 0.0123 (IQR, 0.0068–0.0242) and the median Morisita–Horn index was 0.0257(IQR, 0.0196–0.0493). Raw and standardized cohort medians were closely aligned, with biopsy clonotype sharing changing from 9.1% to 9.2% and shared biopsy read abundance remaining approximately 20.0%. This cohort-level agreement did not imply that individual estimates were unaffected by downsampling; patient-specific raw-versus-standardized comparisons are shown in Supplementary Figure S2, with the corresponding numerical summaries provided in Supplementary Data 4.

The greater contribution of shared clonotypes to biopsy read abundance prompted an examination of peripheral-blood detection across the intratumoral abundance distribution. Biopsy clonotypes were ranked by read abundance and grouped into non-overlapping percentile strata. After matched-depth standardization, median PBMC detection declined across these strata, from 35.9% among the most abundant 1% of biopsy clonotypes to 18.2% in the 1–5% stratum, 15.7% in the 5–10% stratum, 11.9% in the 10–25% stratum, 8.4% in the 25–50% stratum, and 7.0% in the least-abundant half of the repertoire (Figure 2B). Percentile boundaries refer to the number of distinct clonotypes ranked by biopsy abundance, rather than to cumulative fractions of reads. The gradient was directionally consistent within patients, with a negative rank correlation between stratum and PBMC detection in 12 of 13 patients (median Spearman ρ = −0.83), although a strictly monotonic decline across all six strata was observed in five.

This contrast was also evident within patients: the PBMC detection fraction among the most abundant 1% of biopsy clonotypes exceeded that among the least-abundant 50% in 12 of 13 pairs, both before and after depth matching (two-sided Wilcoxon signed-rank P = 0.0005 in both cases; Figure 2C). Thus, the cohort-level abundance gradient was accompanied by a consistent within-patient difference between the two ends of the biopsy abundance distribution. Its persistence after pair-specific depth matching indicates that the pattern was retained under balanced read sampling within pairs, although this procedure does not eliminate differences in sampling completeness or other technical sources of variation between patients.

Because this gradient is a property of individual clonotypes rather than of patient-level summaries, we modelled peripheral detection directly at the clonotype level. Across 62,170 biopsy clonotypes, the odds that a clonotype was also detected in matched blood increased 2.44-fold for each tenfold increase in its intratumoral frequency (95% CI, 2.12–2.80), estimated within patients using patient fixed effects with a cluster-robust variance across the 13 patients. Fitted separately in each patient, the slope was positive in 12 of 13, with a median odds ratio of 2.33 per tenfold increase (Wilcoxon signed-rank P = 0.0007); the single exception was the patient with the lowest-depth biopsy. Peripheral detectability of an intratumoral clonotype is therefore a graded function of its local abundance rather than a property of an arbitrarily defined dominant fraction (Figure 2D).

Shared clonotypes were not merely present in both compartments but were systematically more abundant in the tumor. Among clonotypes detected in both, a median of 66.7% were at least twofold more frequent in the biopsy than in matched blood (range, 21.2–93.5%), compared with a median of 8.7% that were at least twofold more frequent in blood (range, 0–41.0%); the tumor-enriched fraction exceeded the blood-enriched fraction in 12 of 13 patients (Wilcoxon signed-rank P = 0.0007). The median within-patient log2 tumor-to-blood frequency ratio among shared clonotypes was 1.83, corresponding to an approximately 3.6-fold enrichment in the tumor. Shared clonotypes showed a marked tumor-to-blood relative-frequency asymmetry, consistent with intratumoral enrichment. Local expansion is one plausible mechanism, although selective recruitment or retention and compositional effects cannot be excluded. (Figure 2E).

In a separate comparison across patients, exact sequence sharing between biopsies was limited. Of 60,916 distinct biopsy CDR3 amino-acid sequences observed across the cohort, 59,852 (98.3%) were detected in only one biopsy and 1,064 (1.7%) in at least two. Because cohort-wide recurrence is strongly depth-dependent, we repeated this analysis with all biopsies downsampled to the cohort-minimum biopsy depth of 6,194 reads: a median of 22,916 distinct sequences remained, of which 262 (1.1%) were detected in at least two biopsies, so the predominance of patient-private sequences was preserved under this extreme common-depth standardization. Pairwise overlap between biopsies from different patients was correspondingly low across the 78 biopsy pairs, with a median Jaccard index of 0.0015 and a median Morisita–Horn index of 0.0001. These data are summarized in Supplementary Data 5.

Taken together, these observations describe largely individual biopsy repertoires whose overlap with matched blood involves a minority of distinct clonotypes. Within that minority, peripheral detectability showed a graded positive association with intratumoral abundance, and the shared clonotypes themselves are preferentially expanded in the tumor.

### TMB is inversely associated with intratumoral TCRβ effective diversity across the non-hypermutated range

Before testing immune–genomic associations, we assembled an audited subject-level dataset linking paired TCRβ repertoire measurements with OCA Plus molecular data. All 13 curated patients had complete paired PBMC and tumor-biopsy TCR data and complete OCA measurements. All sample-identity, sequencing-depth and clonotype-count consistency checks were passed (Supplementary Data 6).

To define a compact immune panel independently of genomic or clinicopathological associations, TCR measurements were organized into four prespecified biological domains: intratumoral effective diversity, clonal dominance, abundance-conditioned tumor-to-blood clonotype recovery, and abundance-weighted tumor–blood similarity (Table 2). Candidate measurements within competitive domains were evaluated using TCR-only properties, including completeness, value resolution, sequencing-depth association, within-domain representativeness, matched-depth resampling precision, and clonotype denominator support where applicable. Leave-one-subject-out (LOSO) re-selection was used to assess the stability of representative choice. The resulting panel comprised Hill D1, Top-10 cumulative read frequency, PBMC detection of the top 5% of biopsy clonotypes ranked by abundance, and Morisita–Horn similarity. Each representative was recovered in all 13 LOSO analyses. The Top-5% recovery measure retained a low-denominator-support review flag because its smallest subject-specific denominator comprised three clonotypes, but its selection remained stable in 13/13 LOSO runs. No pair of frozen immune representatives reached the prespecified high-redundancy threshold |ρ| ≥ 0.80 (Supplementary Figure S3; Supplementary Data 7).

**Table 2.** Frozen immune and genomic descriptor panels. The four immune and four genomic descriptors carried forward into all association testing, with the candidate measures considered in each biological domain and the outcome-blind rule by which one representative was retained. Immune representatives were selected before any outcome variable was examined; where a domain contained several candidates, the representative with the highest leave-one-subject-out selection stability and the weakest association with sequencing depth was retained. Genomic estimands were defined by the assay and were not selected competitively. Panel composition was frozen before association testing and not revised thereafter.

| Panel | Biological domain | Candidate measures considered | Frozen representative | Selection and audit | Status |
|---|---|---|---|---|---|
| Immune | Intratumoral effective diversity | Hill D1 | Hill D1 | 13/13 LOSO; no review flag | Primary-ready |
| Immune | Intratumoral clonal dominance / concentration | Hill D2; Top-10 frequency; Top-100 frequency; Gini frequency; Fraction of clonotypes to 50% reads | Top-10 frequency | 13/13 LOSO; no review flag | Primary-ready |
| Immune | Abundance-conditioned tumor-to-blood clonotype recovery | Top-1% PBMC recovery; Top-5% PBMC recovery; Top-10% PBMC recovery | Top-5% PBMC recovery | 13/13 LOSO; low denominator support | Primary-ready, review flag |
| Immune | Abundance-weighted tumor-blood similarity | Morisita–Horn | Morisita–Horn | 13/13 LOSO; no review flag | Primary-ready |
| Genomic | YMB | Source-defined continuous assay estimand | TMB | n=13; unique=10; no audit flag | Primary-ready (continuous) |
| Genomic | MSI score | Source-defined continuous assay estimand | MSI score | n=13; unique=13; no audit flag | Primary-ready (continuous) |
| Genomic | Genomic Instability Metric (GIM) | Source-defined continuous assay estimand | GIM | n=13; unique=5; audit flag: high zero mass | Primary-ready (continuous) |
| Genomic | Genomic loss of heterozygosity | Source-defined continuous assay estimand | LOH | n=13; unique=8; no audit flag | Primary-ready (continuous) |

In parallel, four source-defined continuous OCA Plus measures, TMB, MSI score, genomic instability metric (GIM), and loss of heterozygosity (LOH), were frozen without reference to immune associations (Table 2). All four were complete in the 13-patient cohort and satisfied the prespecified technical eligibility criteria. GIM showed a high zero mass, with 9 of 13 tumors having a value of zero and only five distinct values across the cohort; it satisfied the prespecified technical eligibility criteria and was retained, but its limited rank resolution constrains the interpretation of any GIM association. Correlations among the four genomic measures were audited rather than used for data-driven pruning; the largest was between GIM and LOH ( 0.769, below the prespecified high-correlation threshold of 0.80, although high enough that GIM and LOH should not be interpreted as independent genomic dimensions (Supplementary Figure S4; Supplementary Data 8). The frozen four-feature immune and four-feature genomic panels therefore defined 16 prespecified primary comparisons.

Across this primary family, higher TMB was associated with lower intratumoral Hill D1 (Spearman *ρ* = -0.833; two-sided permutation *P* = 0.00072; Figure 3A–B). This was the only association supported by both prespecified multiplicity procedures, with BH *q* = 0.0115 across the 16 primary tests and max-*ρ* permutation FWER *P* = 0.00884. The subject-level bootstrap 95% interval was -0.962 to -0.496. The inverse association was preserved in all 13 LOSO analyses, with *ρ* ranging from -0.875 to -0.787 (Figure 3D). Complete primary effect sizes, uncertainty intervals, permutation results, and LOSO summaries are provided in Supplementary Figure S5A and Supplementary Data 9. The association was not attributable to the two hypermutated tumors, both of which were POLE-mutant: it was preserved after excluding them (ρ = -0.874), after excluding all four tumors carrying the assay POLE flag (ρ = -0.919), and after excluding the two biopsies of lowest sequencing depth (ρ = -0.878). It was also independent of biopsy sequencing depth, which was itself unrelated to TMB (ρ = -0.207, P = 0.50) and which left the association

**Figure 3.**
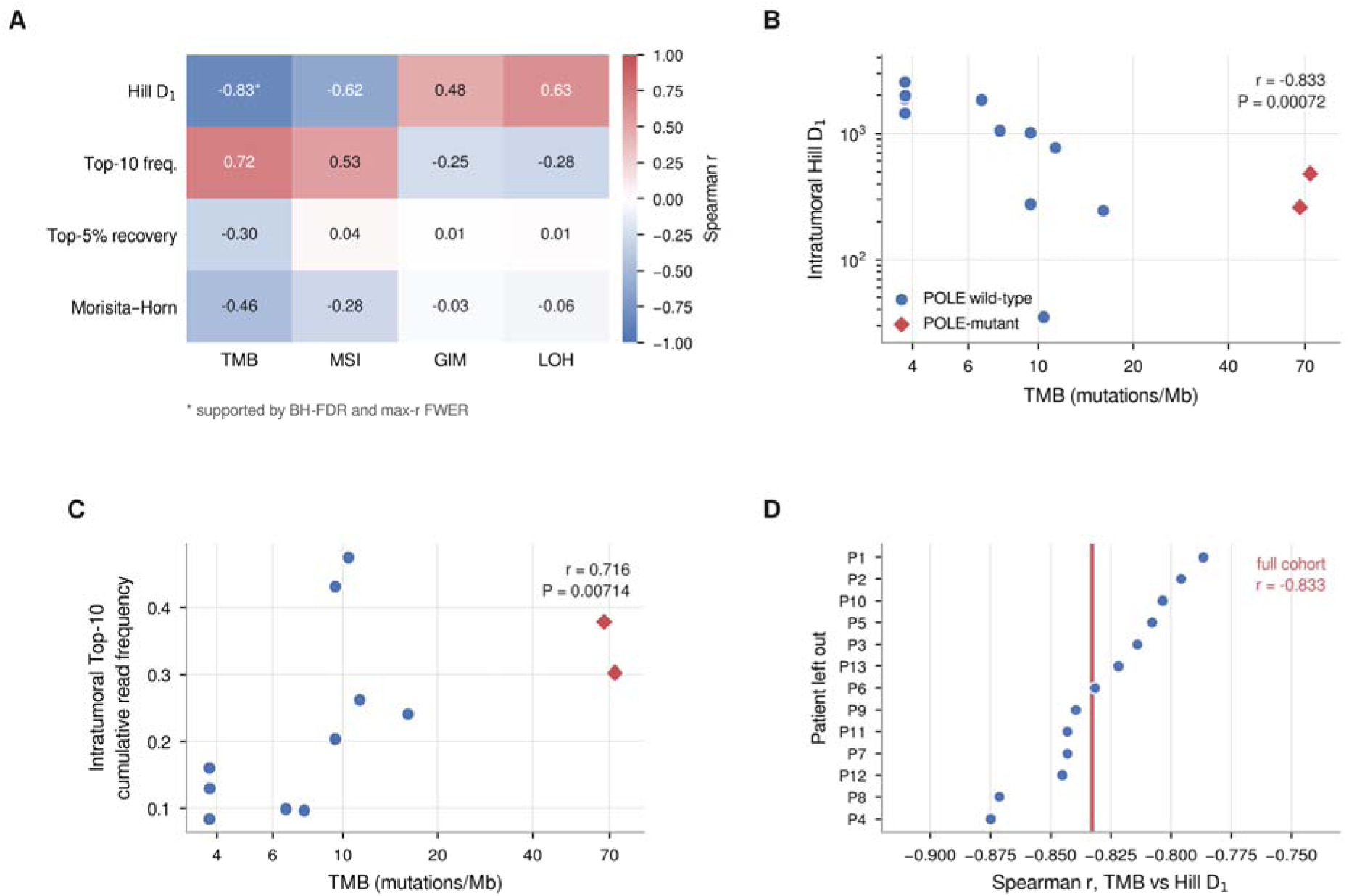
Associations between the frozen immune panel and tumor genomic measures. (A) Spearman correlations for the 16 prespecified immune–genomic comparisons; the asterisk marks the single association supported by both Benjamini–Hochberg false-discovery-rate control and max-r permutation family-wise error control. (B) TMB against intratumoral Hill D_1_ and (C) against intratumoral Top-10 cumulative read frequency; diamonds mark the two POLE-mutant tumors, and both axes in B and the x-axis in C are logarithmic. (D) Leave-one-subject-out values of the TMB–Hill D_1_ correlation, one per patient excluded, with the full-cohort estimate shown as a vertical line.

essentially unchanged in a depth-adjusted partial rank analysis (ρ = -0.831). The relationship was graded rather than driven by hypermutation: among the 11 tumors with TMB below 30 mutations per megabase it was at least as strong as in the full cohort (ρ = -0.874, P = 0.0004), whereas a contrast between the two hypermutated tumors and the remainder was not significant (Mann-Whitney P = 0.31). The four tumors with the lowest TMB values had the four highest Hill D1 values. These TMB measurements were tied at the assay’s lower reporting limit, reducing rank resolution at the low end and warranting cautious interpretation.

TMB also showed a positive association with Top-10 cumulative read frequency (*ρ* = 0.716; permutation *P* = 0.00714; Figure 3C), with a bootstrap 95% interval of 0.319–0.881 and the same direction in all 13 LOSO analyses. However, this association did not meet the prespecified multiple-testing thresholds (BH *q* = 0.0571; max-*ρ* FWER *P* = 0.101). Hill D1 was also positively associated with LOH (*ρ* = 0.630) and negatively associated with MSI score (*ρ* = -0.621), but neither remained supported after multiplicity correction (BH *q* = 0.106 for both). The complete 4 × 4 association structure is shown in Figure 3A and Supplementary Data 9.

The two tumor–blood connectivity descriptors in the primary panel showed no multiplicity-corrected genomic associations. For example, Morisita–Horn similarity showed an inverse association with TMB (r = -0.462), but this was not supported after correction (permutation *P*=0.112, BH *q*=0.256, max-*ρ* FWER *P*=0.750); abundance-conditioned Top-5% PBMC recovery likewise showed no corrected association with TMB, MSI score, GIM, or LOH. A separate 12-test sensitivity family using global biopsy clonotype recovery, Top-10% PBMC recovery, and shared biopsy read abundance yielded no corrected associations (Supplementary Figure S5B; Supplementary Data 10). Exploratory analyses of the four primary immune descriptors according to the OCA POLE source flag also yielded no within-family significant association; the smallest BH-adjusted value was *q* = 0.198 (Supplementary Figure S5C; Supplementary Data 11). Because only two of the four flag-positive tumors carried a pathogenic POLE exonuclease-domain mutation, this flag-based comparison does not correspond to the POLE-mutant molecular class.

Together, these analyses identify an inverse association between TMB and effective intratumoral TCRβ diversity that is graded across the ordinary range of mutational burden rather than confined to hypermutated tumors. A concordant increase in Top-10 clonal concentration was observed with increasing TMB but did not meet the prespecified multiple-testing thresholds, whereas the tested dimensions of tumor–blood repertoire connectivity showed no comparable multiplicity-corrected genomic association.

### Histopathological features show no multiplicity-corrected associations with the frozen immune and genomic panels

We next examined whether the frozen immune and genomic descriptors were associated with conventional histopathological features, without further feature selection. The four immune descriptors and four genomic measures were evaluated against endometrioid grade, myometrial invasion and LVSI, yielding eight prespecified comparisons per endpoint and 24 tests overall. Grade was analyzed ordinally in 11 tumors (7 G1, 3 G2, 1 G3), myometrial invasion in 12 evaluable tumors (8 >50%, 4 ≤50%) and LVSI in 11 (2 positive, 9 negative). Multiplicity was controlled within each eight-test family by Benjamini–Hochberg (BH) correction and by permutation-based max-|effect| family-wise correction, with an additional global BH correction across all 24 comparisons (Supplementary Figure S6; Supplementary Data 12 and 13).

No association remained supported after correction. The two strongest nominal effects were higher LOH in tumors with >50% myometrial invasion (rank-biserial effect = 0.750, exact permutation P = 0.0485) and lower Morisita–Horn tumor–blood similarity with increasing grade (Spearman ρ = −0.576, exact permutation P = 0.0697); neither survived within-family or global correction (minimum BH q = 0.388, minimum max-|effect| FWER P = 0.244, global q = 0.836). For LVSI the smallest nominal P was 0.436. That endpoint had no capacity to yield a corrected association in the first place: with two LVSI-positive tumors the exact label-permutation space contains only 55 assignments, so the smallest attainable two-sided P is 0.036 and the smallest attainable within-family BH q is 0.29, and the negative result therefore reflects the limits of the design rather than evidence against an association. Histotype and p53 pattern were summarized descriptively because of sparse subgroup sizes. Full results for all 24 comparisons are given in Supplementary Data 12–14. None of them showed multiplicity-corrected evidence of association, indicating that the TMB– Hill D1 relationship identified in the immune–genomic analysis was not mirrored by the conventional histopathological features available in this cohort.

## Discussion

This study characterizes the TCRβ repertoire in paired tumor and blood samples from patients with endometrial cancer. Tumor repertoires were narrower than matched peripheral repertoires, as expected for a tissue-infiltrating population, and this difference persisted under all applied depth- and coverage-standardization procedures. However, Hill D2 and Top-10 concentration showed no consistent compartmental shift, indicating that reduced tumor breadth was not equivalent to a generalized increase in clonal dominance. Among biopsy clonotypes, peripheral detectability increased with intratumoral abundance, whereas clonotypes shared between compartments were usually enriched in the tumor. Higher TMB was associated with lower intratumoral Hill D1, whereas tumor–blood connectivity and the evaluated histopathological features showed no multiplicity-corrected associations.

Only a minority of distinct biopsy clonotypes was detectable in matched blood, but their detection followed a graded abundance relationship rather than being restricted to an arbitrarily defined dominant subset. In the clonotype-level model of the observed repertoires, the odds of peripheral detection increased approximately 2.4-fold per tenfold increase in intratumoral frequency. Consistently, the separate matched-depth stratum analysis retained an overall abundance gradient, and patient-specific slopes were positive in 12 of 13 patients. Shared clonotypes were enriched in the tumor by a median of approximately 3.6-fold. Exact CDR3 sharing cannot establish trafficking direction or antigen specificity, and the cross-sectional design cannot determine where proliferation occurred; these findings therefore support intratumoral relative-frequency enrichment consistent with, but not demonstrative of, local expansion.

These findings extend previous TCRβ studies in endometrial cancer. The first linked intratumoral clonality to immune gene expression and progression-free survival but examined tumor tissue only^14^. Subsequent paired tumor–blood studies showed that clonotypes present at multiple tumor sites are over-represented in blood^15^, that circulating tumor-matched clonotypes increase in a subset of pembrolizumab responders^16^, and that such clonotypes may persist for years after treatment of POLE-mutant tumors^17^. Pan-cancer data similarly indicate that tumor-expanded clonotypes can be detected in the circulation^18^. The present study adds a quantitative within-patient analysis showing that peripheral representation tracks intratumoral abundance in a single biopsy. It also shows that, among shared clonotypes, tumor-enriched sequences outnumbered blood-enriched sequences in 12 of 13 patients. This pattern is not readily explained by passive blood carryover alone, although selective recruitment, retention, compositional effects, and local proliferation cannot be distinguished. Technical differences from the study by Kim et al., including RNA rather than genomic DNA, no replicate-concordance filter, retention of all productive clonotypes, and explicit depth resampling, should also be considered when comparing results^15^.

Bulk TCRβ sequencing provides no phenotypic or specificity information. Single-cell studies in other cancers suggest that exhausted intratumoral clones, which are often enriched for tumor reactivity, are less frequently detected in blood than memory-phenotype clones^22^. The shared, tumor-enriched clonotypes observed here may therefore include memory-like tumor-specific cells, bystander clones of unrelated specificity, or both. Any future blood-based application will require paired phenotyping or direct specificity testing to distinguish these possibilities.

The TMB–Hill D1 relationship was the only immune–genomic association supported by both false-discovery-rate and family-wise error correction. Its direction was preserved in every LOSO analysis and remained at least as strong after excluding the two hypermutated POLE-mutant tumors or all tumors carrying the assay POLE flag. This supports a graded relationship that was not driven solely by hypermutated cases. The association is biologically plausible because hypermutated endometrial cancers can generate more candidate neoantigens and often show prominent lymphocytic infiltration^4^, potentially promoting expansion of selected T-cell populations and reducing effective diversity. TMB is not, however, a direct measure of immune recognition, which also depends on neoantigen quality, clonality, expression, and presentation^8,9^. Repertoire architecture may therefore capture a downstream functional dimension of the immune response that is not represented by mutation count alone.

Tumor–blood connectivity and conventional histopathology behaved differently. Neither abundance-conditioned peripheral detectability nor Morisita–Horn similarity showed a genomic association that survived correction, suggesting that the observed genomic signal concerned intratumoral repertoire organization rather than the extent of peripheral representation. Likewise, grade, myometrial invasion, and LVSI showed no corrected association with the selected immune and genomic descriptors. These negative findings do not establish the absence of an association, particularly given the small and imbalanced subgroups, but they are compatible with immune heterogeneity extending beyond the histopathological variables evaluated in this cohort^10^.

These observations may be relevant to immunotherapy, although treatment response was not evaluated. Immune checkpoint inhibitors are now central to the management of advanced and recurrent endometrial cancer^23–26^, yet response heterogeneity remains substantial. Because integrated genomic–immune models have outperformed TMB alone in pan-cancer analyses^13^, repertoire-derived measures could provide complementary information by directly quantifying the organization of the adaptive immune response.

Several limitations should be acknowledged. The cohort was small, limiting statistical precision and representation of molecular subgroups, and the association analyses were exploratory. With only two LVSI-positive tumors, the exact permutation space could not yield a multiplicity-corrected result, so that negative finding primarily reflects the study design. Molecular classes were assigned from sequencing and MSI testing rather than mismatch-repair immunohistochemistry, and subgroup sizes precluded formal comparisons among molecular classes. The cross-sectional design does not address temporal evolution, prognosis, treatment response, trafficking direction, or the site of clonal proliferation. Depth and coverage standardization cannot eliminate differences in tissue cellularity, RNA recovery, or spatial heterogeneity. Clonotypes were defined by CDR3 amino-acid sequence without V or J gene information, potentially merging distinct rearrangements, and abundance was measured using sequencing reads rather than unique molecular identifiers. Finally, resampling, permutation inference, and outcome-blind feature selection improve internal robustness but cannot replace validation in larger independent cohorts.

In conclusion, paired TCRβ profiling revealed a consistent relationship between intratumoral abundance and peripheral detection, together with relative tumor enrichment of shared clonotypes. Higher TMB was associated with lower effective intratumoral diversity, whereas tumor–blood connectivity and the evaluated histopathological features showed no comparable corrected associations. These findings are hypothesis-generating and warrant validation in larger longitudinal studies before any biomarker application.

## METHODS

### Study design and patient selection

This retrospective, observational, cross-sectional exploratory study included 13 women with histologically confirmed endometrial cancer who underwent surgical treatment at the Unit of Obstetrics and Gynaecology of the “Renato Dulbecco” University Hospital in Catanzaro, Italy, between November 2023 and November 2025. The study was non-interventional and involved no investigational drug or medical device.

Patients were eligible for inclusion if they had a histologically confirmed diagnosis of endometrial carcinoma, underwent surgery with availability of tumor tissue suitable for molecular analyses, and had a paired peripheral-blood sample available for TCRβ repertoire analysis. Peripheral blood was collected on the day of surgery as part of routine clinical care, and residual material from samples submitted to the clinical laboratory was used for research analyses. Thus, tumor and peripheral-blood samples represented contemporaneous sampling of the two compartments.

The final study population comprised 13 patients, with a median age of 65 years (interquartile range, 61–70 years). Histopathological characteristics, including histological subtype, tumor grade, depth of myometrial invasion, and lymphovascular space invasion (LVSI), were retrieved retrospectively from pathology records when available. p53 immunohistochemical pattern and regional nodal findings were available only for a subset of patients. Missingness reflected the availability of clinical records and examinations rather than patient selection: the definitive surgical pathology report could not be retrieved for one patient, for whom grade, myometrial invasion, LVSI, and p53 immunohistochemical pattern were left missing rather than inferred. p53 immunohistochemistry had been performed in 7 of 13 patients, and regional nodal findings were documented in 8 of 13. Patient-level clinicopathological data are provided in Supplementary Data 1. The study was approved by the Regional Ethical Review Board of Regione Calabria (Ethics Committee approval no. 84/2026) and was conducted in accordance with the Declaration of Helsinki and Regulation (EU) 2016/679 (General Data Protection Regulation, GDPR). Data and biological samples used for research were pseudonymized using internal laboratory codes. The key linking these codes to patient identities was retained exclusively at the clinical center and was neither shared outside the institution nor deposited in public repositories. All participants provided written informed consent authorizing the use of their biological samples and associated clinical data for research purposes.

### Sample processing and TCRβ sequencing

Peripheral venous blood was collected in EDTA tubes, and peripheral blood mononuclear cells (PBMCs) were isolated by density-gradient centrifugation using Ficoll-Paque (Merck KGaA, Darmstadt, Germany) according to the manufacturer’s instructions. Total RNA was extracted from isolated PBMCs and used for TCRβ repertoire analysis. Fresh tumor specimens were obtained at the time of surgery, and DNA and RNA were extracted from endometrial cancer tissue samples following standardized protocols for downstream molecular analyses^29^.

TCRβ libraries were generated using the Oncomine™ TCR Beta-LR Assay (Thermo Fisher Scientific, Waltham, MA, USA) as previously described. Libraries were barcoded and processed on the Ion Chef™ System (Thermo Fisher Scientific) for template preparation, enrichment, and chip loading onto Ion 530™ chips, followed by sequencing on the Ion GeneStudio™ S5 System (Thermo Fisher Scientific), as previously described^29–31^.

Raw sequencing data were analyzed using MiXCR (version 4.1.2)^32^ with the manufacturer-specific preset optimized for the Oncomine™ TCR Beta-LR Assay. Downstream analyses were conducted on productive TCRβ clonotypes and included estimation of repertoire diversity, clonality, and tumor–blood repertoire overlap metrics^29^.

### Comprehensive genomic profiling

Comprehensive genomic profiling of endometrial tumor samples was performed using the Oncomine™ Comprehensive Assay Plus (Thermo Fisher Scientific), an amplicon-based next-generation sequencing panel designed for simultaneous analysis of DNA and RNA targets^33,34^. The assay interrogates 517 cancer-related genes and supports the detection of single-nucleotide variants (SNVs), insertions/deletions (indels), copy number variations (CNVs), gene fusions, and splice variants, together with the assessment of TMB, MSI, and GIM^33,35^. Libraries were prepared according to the manufacturer’s instructions and sequenced on the Ion GeneStudio™ S5 platform. Sequencing data were processed using the Oncomine™ Comprehensive Assay Plus analysis workflow. TMB was expressed as mutations per megabase, and MSI was assessed using targeted microsatellite loci and summarized by the assay-derived MSI score^33^. Genomic instability was quantified using the GIM, a proprietary score ranging from 0 to 100 that summarizes unbalanced autosomal copy-number changes identified through genomic segmentation^33,35^. The resulting genomic variables, including TMB, MSI score, GIM and LOH, were integrated with TCR repertoire features for downstream immunogenomic analyses.

### Molecular classification

Tumors were assigned to surrogate molecular classes with the hierarchical assay-based algorithm of the WHO classification of female genital tumours: POLE-mutant (POLEmut), then mismatch-repair-deficient (MMRd), then p53-abnormal (p53abn), then no specific molecular profile (NSMP)^36,37^. POLEmut status was defined by a pathogenic POLE exonuclease-domain mutation^38^; the categorical POLE flag reported by the assay, which also captures variants outside the exonuclease domain and copy-number losses, was not used for classification. Because mismatch-repair immunohistochemistry was not available, MMRd status was defined by an MSI-High call of the assay MSI algorithm. p53abn status was defined by a pathogenic or likely pathogenic TP53 variant. POLE and TP53 variants were taken from the unfiltered variant calls of the assay workflow (hg19) and verified independently from the aligned reads of each tumor, by counting alleles at every covered position of the POLE exonuclease domain and the TP53 coding sequence (base quality ≥10, mapping quality ≥4, at least 20 variant reads on both strands, variant allele fraction ≥5%); read-level verification was possible for 12 of 13 tumors, and all variant calls were concordant. Tumors with more than one classifying alteration were assigned according to the hierarchy.

### TCRβ repertoire characterization

Read-based sampling descriptors and repertoire-architecture measures were calculated using custom Python scripts, available on GitHub (https://github.com/cpalmieri71/endometrial-TCRb-repertoire). Within each sample, identical CDR3 amino acid sequences were collapsed by summing read counts, and clonotype relative abundances were calculated by normalizing these counts to the total retained reads. Sampling descriptors included sequencing depth, singleton and doubleton counts, Good’s coverage, and Chao1-estimated richness. Repertoire architecture was characterized by observed clonotype richness (Hill diversity of order 0), Hill diversity of orders 1 and 2 (the exponential of Shannon entropy and inverse Simpson concentration, respectively), the Gini coefficient, the cumulative relative abundance of the 10 and 100 most abundant clonotypes, and the minimum fraction of observed clonotypes required to account for at least 50% of reads. All measures were calculated at the observed sequencing depth, without rarefaction, using read counts rather than cell counts as the abundance measure. The GitHub repository provides detailed documentation of input processing, implemented calculations, mathematical formulas, and output files.

Clonotypes were identified by CDR3 amino acid sequence alone, without reference to V or J gene assignment; sequences sharing a CDR3 across different V–J rearrangements were therefore merged. Abundances were quantified as sequencing read counts rather than unique molecular identifiers, so residual amplification bias is expected to affect measures weighted towards abundant clonotypes (D_2_, Top-10 and Top-100 frequencies) more than observed richness or D_1_. All retained sequences consisted exclusively of the 20 standard amino acids. A minority departed from the expected CDR3 termini or length, taken as a cysteine at the first position, a phenylalanine or tryptophan at the last, and a length of 8–30 amino acids; these accounted for a median of 1.6% of biopsy and 0.7% of PBMC clonotypes and for less than 0.2% of reads in both compartments, and were retained.

Complete definitions, formulas and implementation details for every repertoire measure are given in Supplementary Methods.

### Exact clonotype sharing and sequencing-depth standardization

Exact TCRβ clonotype sharing was analyzed using custom Python scripts, available on GitHub (https://github.com/cpalmieri71/endometrial-TCRb-repertoire). For each matched tumor-biopsy– PBMC pair, we calculated the number of shared clonotypes, the compartment-specific fractions of clonotypes and reads attributable to shared sequences, the Jaccard index, the overlap coefficient, and Morisita–Horn similarity^39^. PBMC detection of biopsy clonotypes was evaluated across six non-overlapping abundance-rank strata (top 1%, 1–5%, 5–10%, 10–25%, 25–50%, and 50–100%) and cumulative top fractions (1%, 5%, 10%, 25%, and 50%). These percentages refer to the number of distinct clonotypes ranked by decreasing biopsy read abundance, rather than their cumulative read contribution. Relative enrichment among shared clonotypes was described using log biopsy-to-PBMC relative-frequency ratios.

Sensitivity to sequencing depth was assessed using 500 repeated downsampling iterations without replacement, implemented by multivariate hypergeometric sampling to the lower read depth of each pair. Overlap and detection measures were recalculated at each iteration, including biopsy abundance ranks and stratum membership, and summarized by resampling medians and 2.5th–97.5th percentile ranges. These ranges describe conditional read-sampling variability, not biological confidence intervals.

Pairwise overlap was also calculated between biopsies from different patients, with pair-specific depth matching. Cross-patient clonotype recurrence (*publicness*) was described by the number of biopsies containing each sequence and the fractions of clonotypes and reads attributable to sequences detected in at least two or three patients. Observed publicness measures were retained as the primary descriptors; downsampling all biopsies to the cohort-wide minimum depth provided an additional sensitivity analysis. These analyses were descriptive. The script and detailed documentation of input processing, mathematical formulas, resampling procedures, and output files are available on GitHub (https://github.com/cpalmieri71/endometrial-TCRb-repertoire).

Complete definitions and formulas for the overlap measures, the matched-depth resampling procedure, the abundance-rank strata and the between-patient recurrence analysis are given in Supplementary Methods.

### Clonotype-level modelling of peripheral detection

Peripheral detectability was additionally modelled at the level of individual clonotypes. For each biopsy clonotype, detection in the matched PBMC repertoire was treated as a binary outcome and modelled by logistic regression on the base-10 logarithm of its intratumoral frequency, with patient fixed effects so that the coefficient is identified within patients, and a cluster-robust sandwich variance estimator over the 13 patients. The same model was fitted separately within each patient, and the resulting slopes were compared with zero using the two-sided Wilcoxon signed-rank test; this patient-level test, rather than the pooled model, provides the inference that respects the number of independent subjects. Relative abundance between compartments was summarized for clonotypes detected in both, as the fraction at least twofold more frequent in each compartment and as the median within-patient log_2_ tumor-to-blood frequency ratio.

### Outcome-blind construction of the immune and genomic panels

Candidate immune descriptors were grouped into four prespecified biological domains, intratumoral effective diversity, intratumoral clonal dominance, abundance-conditioned tumor-to-blood recovery, and tumor–blood abundance similarity, and a single representative measurement was selected within each domain. Selection used repertoire and overlap data only: genomic, clinical and histopathological variables were excluded programmatically, and the selection script terminates with an error if any such variable enters candidate scoring. Candidates were screened for completeness, variability, residual association with sequencing depth and within-domain redundancy, and the stability of each selection was assessed by repeating the entire procedure with each subject omitted in turn. Where a domain offered several abundance thresholds, the competing thresholds were compared on the same outcome-blind criteria.

For downstream primary immunogenomic analyses, the frozen immune panel comprised intratumoral Hill, intratumoral top-10 cumulative read frequency, the depth-standardized PBMC detection fraction of the most abundant 5% of biopsy clonotypes, and depth-standardized Morisita– Horn similarity. These descriptors represented effective diversity, clonal dominance, abundance-conditioned tumor-to-blood recovery, and abundance-weighted tumor–blood similarity, respectively. They were retained as individual variables rather than combined into composite focusing or sharing scores, and were evaluated against four separately defined continuous genomic measures: TMB, MSI score, genomic instability metric, and loss of heterozygosity. Feature selection was performed independently of immune–genomic association results. The primary TMB association was additionally examined in post hoc sensitivity analyses that excluded the two hypermutated tumors, all tumors carrying the assay POLE flag, and the two biopsies of lowest sequencing depth, and in a partial rank analysis adjusting for biopsy sequencing depth.

Four continuous genomic measures, TMB, MSI score, genomic instability metric and loss of heterozygosity, were frozen in parallel from the Oncomine Comprehensive Assay Plus output without reference to immune data. Pairwise correlations among the selected immune representatives and among the genomic measures were audited against a prespecified redundancy threshold of |r| ≥ 0.80 but were not used for further data-driven pruning.

### Statistical analysis

Paired comparisons between compartments used the two-sided Wilcoxon signed-rank test across the 13 matched pairs. Monotonic associations between continuous variables were quantified by the Spearman rank correlation coefficient, and associations with binary endpoints by the rank-biserial correlation.

Because observed richness and D_1_ are not depth-corrected, all compartmental comparisons were repeated under three standardizations. First, within each pair the deeper repertoire was randomly subsampled without replacement to the depth of the shallower member, with 100 subsamples per repertoire and metrics averaged across subsamples. Second, all repertoires were rarefied to a cohort-wide common depth of 63,199 reads, retaining the 11 pairs in which both members reached that depth; an intermediate depth of 12,792 reads was also evaluated, retaining 12 pairs. Third, all 26 repertoires were standardized to a common Good’s coverage of 0.838, the highest value attainable in every repertoire, following coverage-based standardization as described by Chao and Jost^40^. Rarefaction curves were computed as exact hypergeometric expectations rather than by simulation. Paired compartments were compared using the two-sided Wilcoxon signed-rank test; P values are reported as descriptive, without correction for multiple comparisons. Results of all standardized analyses are provided in Supplementary Data 3.

Immune–genomic associations were tested as a prespecified family of 16 comparisons between the four frozen immune descriptors and the four frozen genomic measures. Inference was by permutation: subject labels of the genomic block were permuted 100,000 times, with the same permutation applied to all four genomic measures so that their mutual correlation was preserved, and two-sided P values were computed as (k + 1)/(B + 1), where B is the number of permutations and k the number yielding an absolute correlation at least as large as the observed one. Multiplicity was controlled in two complementary ways: Benjamini–Hochberg false-discovery-rate correction within the family, and a permutation-based family-wise error rate derived from the null distribution of the maximum absolute correlation across all 16 comparisons. Uncertainty around each coefficient was summarized by subject-level bootstrap resampling with percentile intervals, and stability by recomputing each coefficient with one subject omitted in turn. A secondary family of 12 recovery-sensitivity comparisons and an exploratory family of four comparisons according to the OCA POLE source flag were analysed separately, each with its own within-family correction.

Associations with histopathological endpoints were analysed as three endpoint families of eight comparisons each: endometrioid grade, treated as ordinal; myometrial invasion exceeding 50% of the myometrial thickness; and LVSI. Because the label spaces were small enough to enumerate exhaustively, permutation P values were obtained from the complete set of distinct label assignments rather than by Monte Carlo sampling. Multiplicity was controlled within each family by Benjamini–Hochberg correction and by a permutation-based maximum-effect family-wise procedure, with an additional global Benjamini–Hochberg correction across all 24 comparisons. The resolution attainable by these tests is bounded by the size of the permutation space: with two LVSI-positive tumors only 55 distinct assignments exist, so the smallest attainable two-sided P value is 0.036 and the smallest attainable within-family q value is 0.29.

P values are reported as descriptive measures of evidence. Multiplicity was controlled within each prespecified family, and additionally across the three histopathological endpoint families; no correction was applied between the immune–genomic and histopathological blocks, nor between the primary, secondary and exploratory association families, and the study was not powered for formal hypothesis testing.

Analyses were performed in Python 3.9.22 with NumPy 2.0.2, pandas 2.3.3, SciPy 1.13.1, matplotlib 3.9.4 and powerlaw 2.0.0; sensitivity analyses, figures and Supplementary Data were generated in Python 3.13 with NumPy, pandas, SciPy, matplotlib and openpyxl; the exact versions are listed in the repository. The complete version-controlled pipeline, software environments, configuration files, random seeds and script checksums are available in the public repository (https://github.com/cpalmieri71/endometrial-TCRb-repertoire).

## Data Availability

All data produced are available online at the NCBI Sequence Read Archive (BioProject PRJNA1474700, study SRP706670; raw TCRβ FASTQ files, https://www.ncbi.nlm.nih.gov/bioproject/PRJNA1474700), Zenodo (processed clonotype tables, https://doi.org/10.5281/zenodo.24139417) and GitHub (analysis code, https://github.com/cpalmieri71/endometrial-TCRb-repertoire). Patient-level clinicopathological and genomic data and all source data underlying the figures are provided in the Supplementary Data.

https://www.ncbi.nlm.nih.gov/bioproject/PRJNA1474700

https://github.com/cpalmieri71/endometrial-TCRb-repertoire

## Data availability

Raw TCRβ sequencing FASTQ files generated during the current study have been deposited in the NCBI Sequence Read Archive (SRA) under BioProject accession PRJNA1474700, “Integrated T-cell Receptor Repertoire and Tumor Immunogenicity Profiling in Endometrial Cancer” (study accession SRP706670). BioSample and run accessions for all 26 repertoires (13 tumor biopsies and 13 matched PBMC samples) are listed in Supplementary Data 15.

Processed per-repertoire clonotype tables generated using the MiXCR pipeline ^32^ and used as input for all analyses reported here are available on Zenodo (https://doi.org/10.5281/zenodo.24139417). Each table lists, for every clonotype, the CDR3 amino acid sequence (aaSeqCDR3), its read count (readCount) and its relative read frequency (readFraction). File names correspond to the patient codes (P1–P13) used in Supplementary Data 1.

Patient-level clinicopathological and genomic data for all 13 patients are provided in Supplementary Data 1. Per-repertoire sampling and architecture metrics and their sensitivity to sampling standardization are provided in Supplementary Data 2 and 3, within-patient tumor–blood overlap, abundance-stratified sharing and between-patient recurrence in Supplementary Data 4 and 5, subject-level integration, panel construction and immune–genomic association outputs in Supplementary Data 6–11, and histopathological association outputs in Supplementary Data 12–14. Together these files contain the source data underlying every figure and statistical analysis reported here.

The analysis code, including the scripts that regenerate every figure and Supplementary Data file from the deposited clonotype tables, is available at https://github.com/cpalmieri71/endometrial-TCRb-repertoire.

## Acknowledgments

This work was funded by the Italian Ministero della Salute Grant RF-2019-12370255 to C.P., G.V., F.Z. and M.G., and Ministero dell’Università e della Ricerca, Grant PNC 0000001 D.4Health to C.P. and G.C.

During the preparation of this work, the author(s) used ChatGPT (OpenAI) to support manuscript language editing, technical documentation, coding assistance, code review, debugging, and graphical rendering. All scientific questions, analytical definitions, estimands, parameter choices, workflow architecture, data interpretation, and final methodological decisions were defined and validated by the authors. AI-assisted code suggestions were inspected, tested, and revised by the authors before inclusion in the production pipeline, and all reported analyses were generated using the finalized, version-controlled codebase.

## Author Contributions

I.A., A.A., A.I., Methodology, Investigation, Writing-Original draft preparation.

C.D.M., M.T.D.A, Methodology.

G.F., Methodology, Supervision.

T.S., R.V., Clinical Sample Collection.

F.M.Z., C.C., Data curation.

C.P., A.A, I.A. Writing-Original draft preparation.

R.G., L.I.R., G.F. Writing-Original draft preparation.

C.G., M.G., G.S. Writing-review and editing.

C.P., F.C., G.V., M.G., F.Z., Funding acquisition, Supervision.

I.A., A.A, and A.I, contributed equally and share first authorship.

C.P., F.Z., and G.V., contributed equally and share senior authorship.

## DECLARATIONS

### Ethical approval

This study is a retrospective study and was approved by the Regional Ethical Review board of Regione Calabria (Ethics committee approval number: 84/2026; Clinical trial number: not applicable). The study was conducted in accordance with the Declaration of Helsinki, and EU Regulation 2016/679 (GDPR). Data were pseudonymized: samples were identified by internal laboratory codes, and the information linking these codes to patient identity was held at the clinical center and was neither shared outside it nor deposited.

### Consent to participate

All participants provided written informed consent authorizing the use of their pseudonymized biological material and associated clinical data for research purposes, at the time of sample collection.

### Consent to publish

Not applicable; no identifying information or images of individual patients are published.

