## Supplementary Figures S1-S6 for "Tumor–blood TCRβ repertoire architecture in endometrial cancer: abundance-dependent peripheral detection and intratumoral enrichment of shared clonotypes"

Aversa I & Abatino A et al.

### SUPPLEMENTARY FIGURES

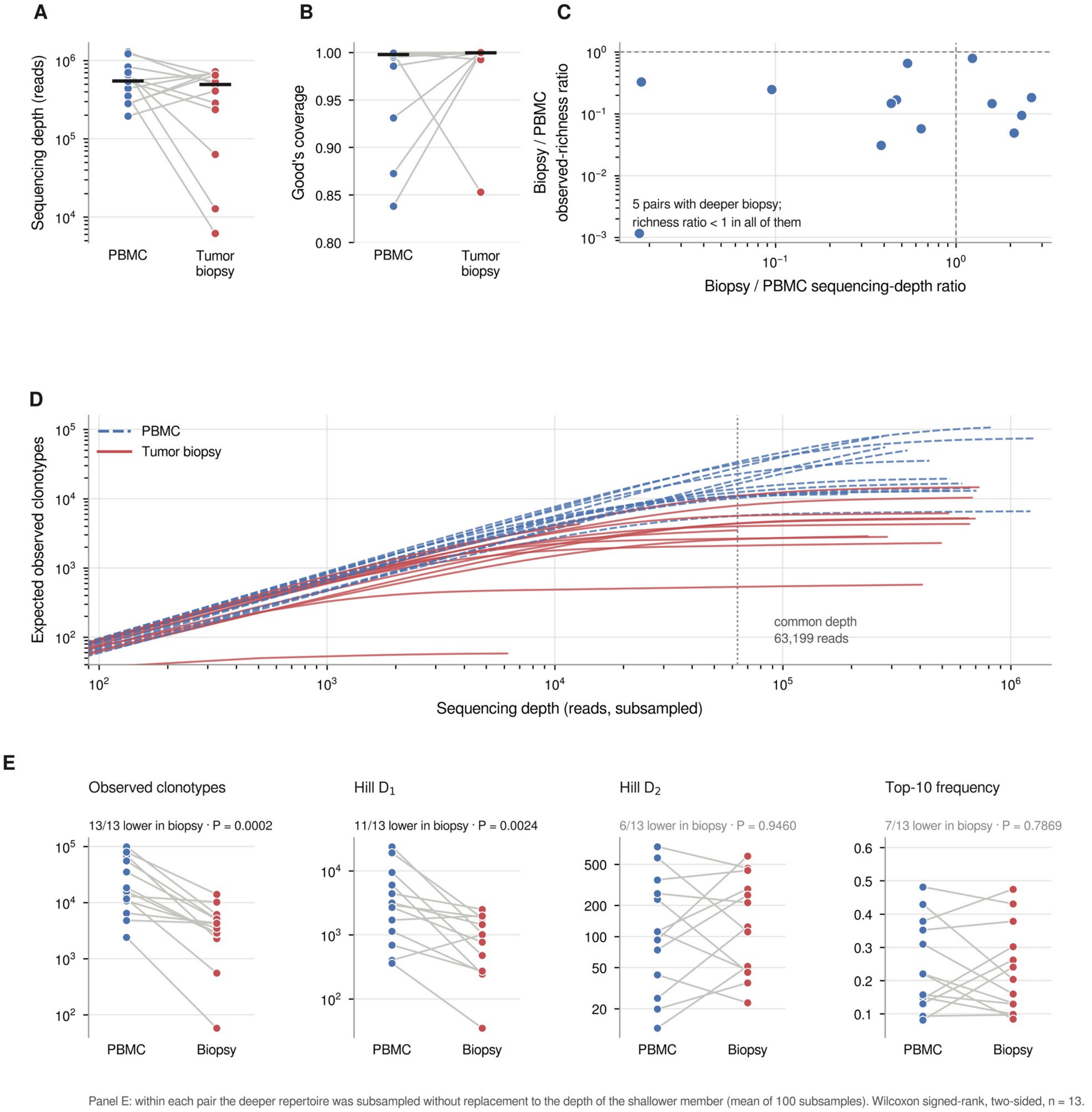


**Supplementary Figure S1.** Sequencing depth, sampling coverage, and depth–richness relationship in paired PBMC and tumor-biopsy TCRβ repertoires, and sensitivity of compartmental differences to sampling standardization. **(A)** Paired productive TCRβ sequencing depth in peripheral-blood mononuclear cell (PBMC) and tumor-biopsy repertoires from 13 patients. Each line connects the matched samples from the same patient;

horizontal black bars indicate cohort medians. The y-axis is logarithmic. **(B)** Good's coverage for paired PBMC and tumor-biopsy repertoires. Lines connect matched samples and horizontal black bars indicate cohort medians. **(C)** Relationship between the biopsy/PBMC sequencing-depth ratio and the biopsy/PBMC observed-richness ratio for each patient. Both axes are logarithmic; dashed reference lines at a ratio of 1 indicate equal sequencing depth and equal observed richness between compartments, respectively. Each point represents one matched biopsy–PBMC pair. (D) Rarefaction curves for all 26 repertoires, computed as exact hypergeometric expectations; the dotted vertical line marks the cohort-wide common depth of 63,199 reads used for the depth-standardized analysis. Both axes are logarithmic. (E) Paired comparisons of observed clonotypes, Hill D1, Hill D2 and Top-10 cumulative read frequency after per-pair rarefaction, in which the deeper repertoire of each pair was subsampled without replacement to the depth of the shallower member. Directional counts and two-sided Wilcoxon signed-rank P values are given above each panel.

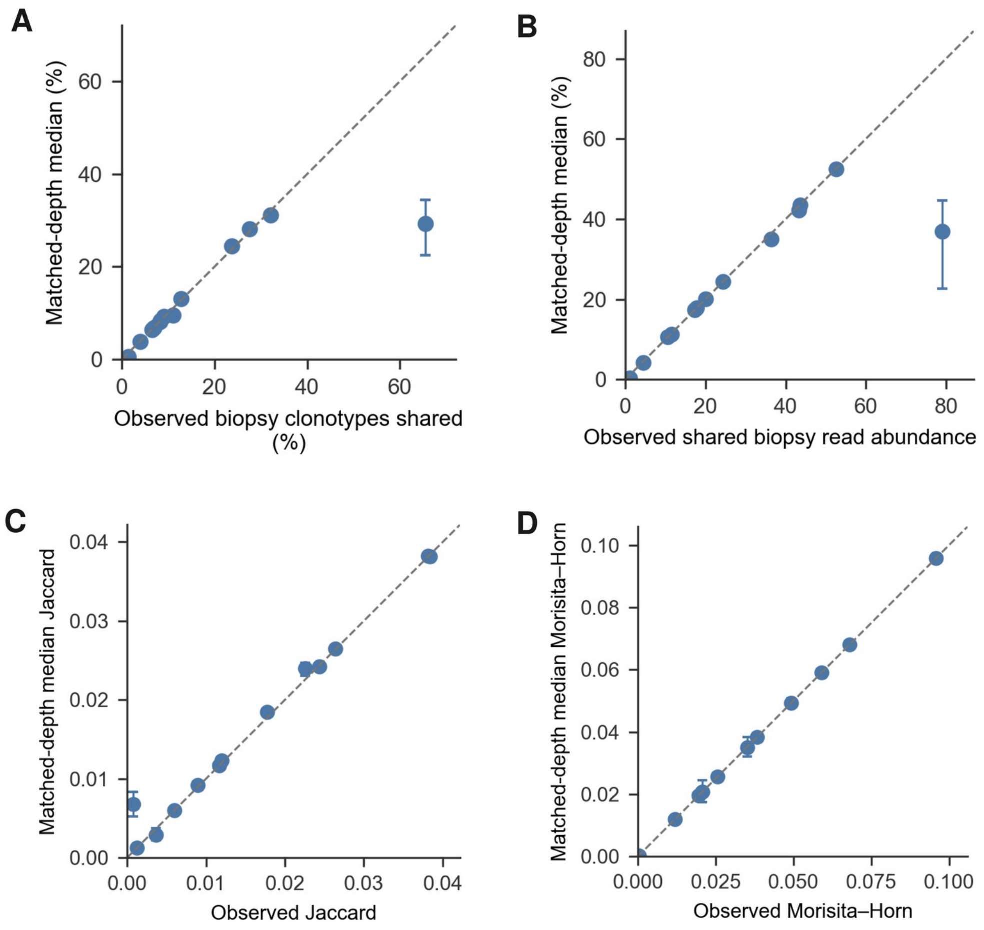


**Supplementary Figure S2. Sensitivity of paired tumor–blood sharing metrics to matched-depth standardization.**

**(A)** Observed fraction of biopsy clonotypes detected in matched PBMC versus the corresponding matched-depth median. **(B)** Observed fraction of biopsy read abundance contributed by clonotypes also detected in matched PBMC versus the corresponding matched-depth median. **(C)** Observed Jaccard index versus matched-depth median Jaccard index. **(D)**Observed Morisita–Horn index versus matched-depth median Morisita–Horn index. Each point represents one patient. Matched-depth values are patient-specific medians across 500 downsampling replicates to the lower sequencing depth of each biopsy–PBMC pair. Vertical error bars indicate the 2.5th–97.5th percentile range across downsampling replicates. Dashed diagonal lines indicate equality between observed and matched-depth values.

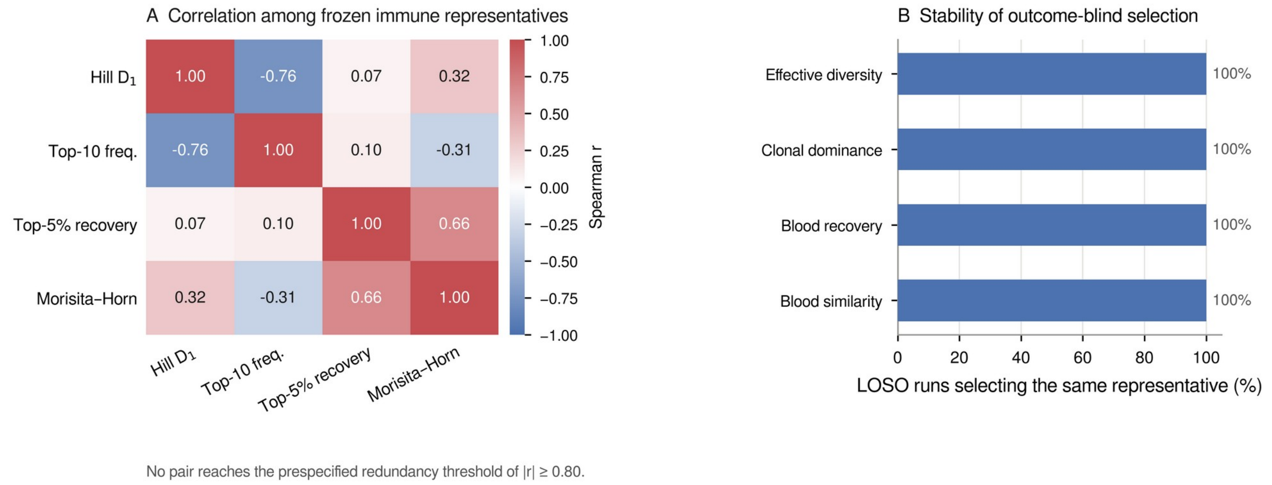


**Supplementary Figure S3.** Outcome-blind construction of the frozen immune panel. (A) Spearman correlations among the four frozen immune representatives; no pair reaches the prespecified redundancy threshold of $|r| \geq 0.80$. (B) Percentage of leave-one-subject-out runs in which each biological domain selected the same representative measurement.

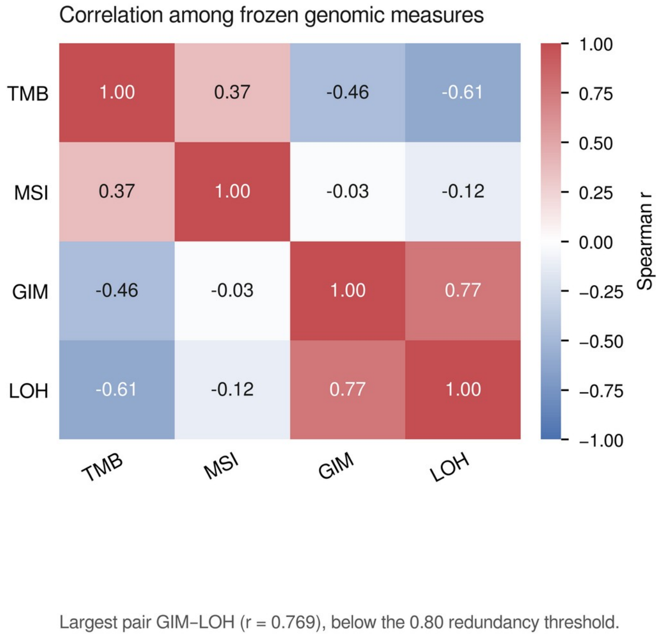


**Supplementary Figure S4.** Spearman correlations among the four frozen genomic measures. The largest pairwise correlation, between the genomic instability metric and loss of heterozygosity (r = 0.769), lies below the prespecified redundancy threshold of 0.80 but indicates that these two measures are not independent genomic dimensions.

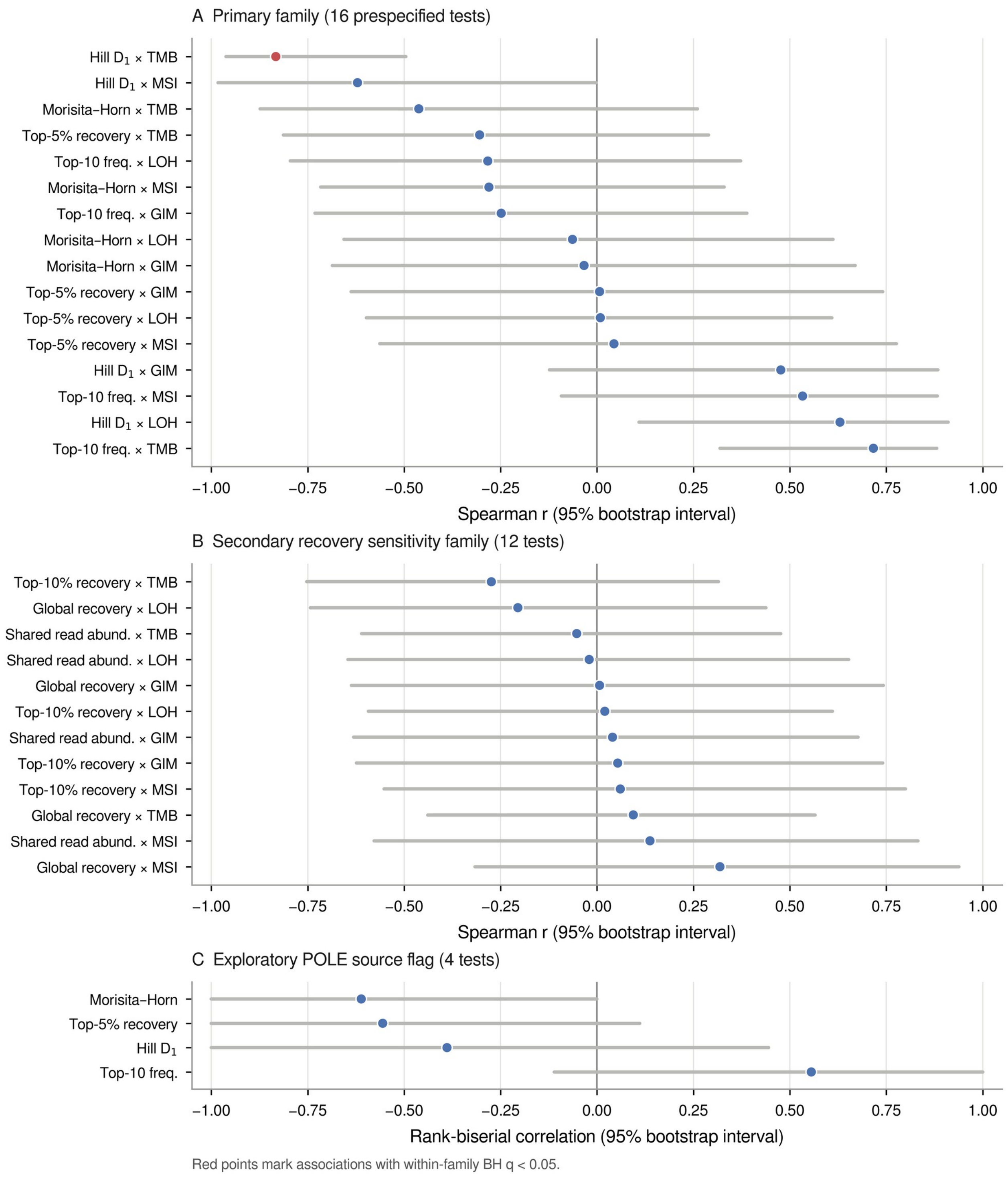


**Supplementary Figure S5.** Effect sizes and uncertainty for the immune–genomic association families. (A) The 16 prespecified primary comparisons, (B) the 12-test secondary recovery sensitivity family, and (C) the four exploratory comparisons according to the OCA POLE source flag. Points are Spearman correlations, or rank-biserial correlations in C, with subject-level 95% bootstrap percentile intervals; red points mark associations with within-family Benjamini–Hochberg $q < 0.05$. Comparisons are ordered by effect size within each family.

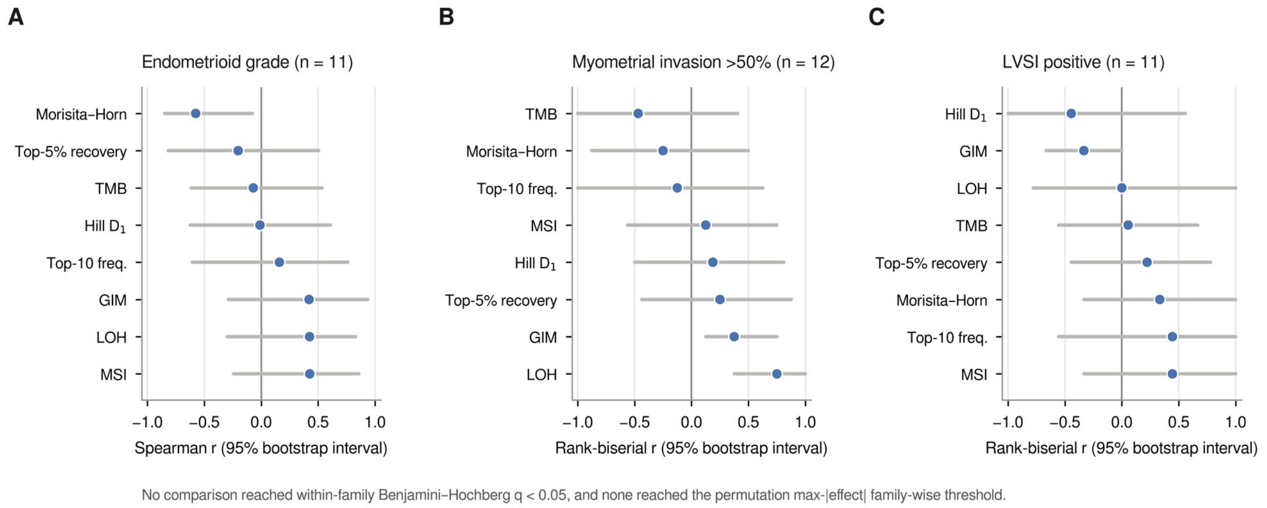


**Supplementary Figure S6**. Frozen immune and genomic descriptors against histopathological endpoints. (A) Endometrioid grade analysed ordinally in 11 tumors; (B) myometrial invasion exceeding 50% of the myometrial thickness, in 12 evaluable tumors; (C) lymphovascular space invasion, in 11 evaluable tumors. Points are Spearman correlations for grade and rank-biserial correlations for the binary endpoints, with subject-level 95% bootstrap percentile intervals. No comparison reached within-family Benjamini–Hochberg $q < 0.05$, and none reached the permutation max-|effect| family-wise threshold. With two LVSI-positive tumors the exact label-permutation space admits no multiplicity-corrected result, so panel C is shown for completeness.
