## Supplementary Methods for "Tumor–blood TCRβ repertoire architecture in endometrial cancer: abundance-dependent peripheral detection and intratumoral enrichment of shared clonotypes"

Aversa I & Abatino A. et al.

### SUPPLEMENTARY METHODS

#### Repertoire metric definitions and formulas

TCRβ repertoires were analyzed using upstream-filtered productive clonotype tables, with exact CDR3 amino acid sequences serving as clonotype identifiers. Within each sample, entries with identical CDR3 amino acid sequences were collapsed by summing their read counts. Clonotype frequencies were calculated as $p_i = c_i/N$, where $c_i$ denotes the retained read count of clonotype $i$ and $N = \sum_i c_i$ is the total retained read count. Tumor-biopsy and peripheral blood mononuclear cell (PBMC) repertoires were treated as distinct, patient-matched biological samples rather than technical replicates. Within-repertoire descriptors were calculated at the observed sample-specific sequencing depth, without rarefaction; depth- and coverage-standardized analyses are described below.

Observed richness ($S_{\text{obs}}$) was defined as the number of distinct CDR3 amino acid sequences. Effective repertoire diversity was summarized using Hill numbers of orders 1 and 2:

$$D_1 = \exp\left(-\sum_i p_i \ln p_i\right), \qquad D_2 = \frac{1}{\sum_i p_i^2}.$$

Here, $D_1$ corresponds to the exponential of Shannon entropy, whereas $D_2$ corresponds to inverse Simpson diversity and places greater weight on abundant clonotypes. Clonal dominance was primarily described by the cumulative read frequency of the ten most abundant clonotypes. Additional descriptive measures included the cumulative frequency of the 100 most abundant clonotypes, the uncorrected Gini coefficient of clonotype frequencies, and the minimum fraction of observed clonotypes required to account for 50% of reads. The latter was calculated by ranking clonotypes in descending abundance, identifying the smallest number whose cumulative frequency reached or exceeded 0.50, and dividing this number by $S_{\text{obs}}$.

Read-sampling characteristics were described using sequencing depth, singleton and doubleton counts, Good's coverage, and the Chao1 richness estimator. Good's coverage was calculated as $1 - f_1/N$, where $f_1$ is the number of clonotypes represented by one read. Chao1 was calculated as:

$$\hat{S}_{\text{Chao1}} = \begin{cases} S_{\text{obs}} + \dfrac{f_1^2}{2f_2}, & f_2 > 0, \\ S_{\text{obs}} + \dfrac{f_1(f_1 - 1)}{2}, & f_2 = 0, \end{cases}$$

where $f_2$ denotes the number of clonotypes represented by two reads. These measures were retained as sampling-sensitive descriptors: singleton and doubleton counts refer to sequencing reads rather than one-cell or two-cell clone sizes, and Good's coverage was not interpreted as the fraction of all biological clonotypes recovered.

To limit redundant representation of repertoire properties, algebraic aliases and jointly derived measures, including separate Shannon entropy, Simpson concentration, Gini–Simpson, Pielou evenness, and normalized Shannon clonality variables, were not retained as additional features in the canonical analysis tables. Repertoire characteristics were summarized by compartment using medians and interquartile ranges.

**Tumor–blood overlap and sequencing-depth standardization**

Within-patient tumor–blood sharing was defined by exact CDR3 amino acid identity. Tumor and PBMC repertoires were aligned over the union of their observed clonotypes, assigning zero counts to sequences absent from either compartment. Set-based measures included the number of shared clonotypes, Jaccard similarity, and the overlap coefficient:

$$J_{\mathrm{T,P}} = \frac{|T \cap P|}{|T \cup P|}, \qquad O_{\mathrm{T,P}} = \frac{|T \cap P|}{\min(|T|, |P|)},$$

where $T$ and $P$ denote the tumor and PBMC clonotype sets, respectively. Directional sharing was quantified separately as the fraction of tumor clonotypes detected in PBMC, $|T \cap P|/|T|$, and the fraction of PBMC clonotypes detected in the tumor, $|T \cap P|/|P|$. For each compartment, the shared read fraction was calculated by summing the frequencies of clonotypes present in both samples. Abundance-weighted similarity was assessed using the Morisita–Horn index:

$$MH_{\mathrm{T,P}} = \frac{2 \sum_i p_{i,T}\, p_{i,P}}{\sum_i p_{i,T}^2 + \sum_i p_{i,P}^2},$$

with frequencies normalized separately within each repertoire.

Raw overlap measurements were retained, and their sensitivity to unequal sequencing depth was assessed using 500 repeated matched-depth resamplings per patient. The target depth was the lower total read count of the paired tumor and PBMC repertoires. Read counts were downsampled without replacement using multivariate hypergeometric sampling; a repertoire already at the target depth was retained unchanged. Overlap measures were recalculated for each realization.

Depth-standardized estimates were summarized by the median across resamplings, with empirical 2.5th–97.5th percentile intervals and coefficients of variation retained to describe resampling variability. These intervals characterize variability under repeated read subsampling rather than between-patient uncertainty. Depth matching was performed within each pair, not at a common depth across all patients, and did not correct for upstream differences in RNA input, T-cell content, extraction, amplification, or library preparation.

**Abundance-conditioned sharing and between-patient recurrence**

To assess whether PBMC detection varied with intratumoral clonotype abundance, biopsy clonotypes were ranked by descending read count and partitioned into non-overlapping percentile strata: the most abundant 1%, followed by the 1–5%, 5–10%, 10–25%, 25–50%, and 50–100% rank intervals. Percentile boundaries referred to the number of distinct clonotypes, not cumulative read abundance. Within each stratum, the PBMC detection fraction was calculated as the proportion of biopsy clonotypes also observed in the paired PBMC sample. Cumulative top-rank subsets comprising the most abundant 1%, 5%, 10%, 25%, and 50% of biopsy clonotypes were evaluated separately. For a cumulative fraction $f$, the subset contained $\max[1, \lceil f S_{\mathrm{obs}} \rceil]$ clonotypes. Abundance ties were resolved deterministically by CDR3 sequence order.

Abundance rankings, stratum membership, and cumulative top-rank subsets were recalculated within each matched-depth realization before computing PBMC detection fractions. Raw and depth-standardized measurements were retained separately. PBMC detection in the most abundant 1% and the least-abundant 50% of biopsy clonotypes was compared across patients using the two-sided Wilcoxon signed-rank test, and the monotonicity of the abundance gradient was summarized per patient by the Spearman rank correlation between stratum order and PBMC detection fraction. P values are reported as descriptive, without correction for multiple comparisons.

Between-patient sharing was assessed by pairwise comparison of biopsy repertoires and by counting the number of patients in whom each exact CDR3 amino acid sequence was detected. Sequences observed in only one biopsy were classified as cohort-private, whereas those detected in at least two biopsies were classified as recurrent or public within the cohort. Pairwise biopsy overlap was also evaluated at pair-specific matched depths. Raw recurrence estimates were retained as the primary descriptive measurements; repeated downsampling of all biopsies to the cohort-wide minimum biopsy depth was used as a separate sensitivity analysis.

**Subject-level data integration**

Repertoire descriptors and tumor–blood overlap measurements were integrated with clinical, histopathological, and tumor-derived Oncomine Comprehensive Assay Plus data in an audited dataset containing one record per patient. The curated clinical cohort defined the subject set. Before integration, sample identities and paired-sample completeness were verified, and sequencing depths, observed clonotype counts, matched-depth targets, and resampling counts were checked for consistency across analytical steps. Previously calculated repertoire and overlap measurements were imported without recomputation. Tumor genomic measurements were linked at the patient level and were not assigned to PBMC samples. Technical provenance and tail-fitting variables were retained in the complete master dataset but excluded from the analysis-ready repertoire feature set.
